# Patterns of Herbal Medicine Use in a General Outpatient Clinic in Nigeria- A Cross-sectional Study

**DOI:** 10.1101/2024.01.30.24302041

**Authors:** Afisulahi Abiodun Maiyegun, Mark Divine Akangoziri, Bukar Alhaji Grema, Yahkub Babatunde Mutalub, Farida Buhari Ibrahim, AbdulRauf Segun Ibraheem

## Abstract

**Background:** Herbal medicine use remains an important part of primary care and the World Health Organization has mandated member countries to conduct research into this and other forms of traditional medicine. However, research into traditional medicine remains scanty, even in the developing where it is often a major health option.

**Objective:** to determine the prevalence, types and sources of herbal medicine used among the study population.

**Methods:** Three hundred and forty-one questionnaires were administered to adult patients attending the general outpatient clinic of a tertiary hospital in Northern Nigeria. The data was collected and analysed using EPI INFO version 7.2.5.0 software.

**Results:** The prevalence of herbal medicine use was 85.34%; the commonest herbs were moringa (59.8%), black seed (36.8%), and olive oil (34.4%); and the commonest source of the herbs were herbal practitioners and herbal medicine vendors (79.04%). Herbal medicine use was associated with religion (P= 0.0005) and residential area (P= 0.01). Only 18.12% of participants ever discussed herbal medicine use with their doctors.

**Conclusion:** herbal medicine use remains high even among patients attending outpatient clinics in tertiary hospitals in Nigeria. However, health workers are often not the source of herbal medicine and patients rarely disclose its use to them.

## Introduction

The World Health Organization (WHO) estimates that up to 80 % of the African population use traditional medicine for their health needs[1]. It also acknowledges the role of traditional medicine in the global aim to achieve universal health coverage.[2] In recognition of this role, the WHO has mandated member states to increase research into traditional medicine used in their regions. This will include research into the types of traditional medicine, the reasons for use, the sources, and the safety of such medicines, among other related issues.[2]

In Nigeria, relatively few studies have addressed the use of traditional medicine among the population – though available local studies suggest a high prevalence – and majority of these studies have been conducted in the southern parts of the country, with very scanty literature about the traditional medical practices in Northern Nigeria.[3–6] Since the country is characterized by significant geographical contrast in flora, culture and religion, factors which drive the use of traditional medicine, studies in Northern Nigeria are required to fill the knowledge gap.

Herbal medicine are the commonest traditional medicines.[7,8] Herbs are also one of the most likely traditional medicines to intersect with orthodox medicine due to the side effects of herbs and drug-herb interactions.[9,10] This study, therefore, aimed to determine the types and characteristics of herbal medicine use among primary care clinic attendees in Bauchi, North-eastern Nigeria.

## Methods

The study was conducted in the general outpatient unit of Abubakar Tafawa Balewa University Teaching Hospital (ATBUTH), Bauchi, Northern Nigeria, between September and December 2018. The study population were adult patients (at least 18 years old) attending the general outpatient clinic. Patients who were very sick or declined consent were excluded from the study. Sample size for the study was determined using the Cochran’s formula: N = Z^2^pq/d^2^, where N = minimum sample size, Z = confidence level at 95% (1.96), p = 0.67, being the prevalence of herbal medicine use from a previous study in Nigeria (Oreagba et al), q = (1-p), and d = level of precision (0.05).[3] The sample size was 341. Prospective participants were selected through systematic random sampling, and their written consent sought and obtained before enrolment. The study was approved by the Ethical Review Committee of ATBUTH (Approval number: 03/03/2018).

An interviewer-administered, semi-structured questionnaire was used for data collection. The questionnaire had five sections: biodata, family characteristics, family income, medical history and herbal medicine use. Data was analyzed using Epi Info version 7.2.5.0 software developed by the Centers for Disease Control and Prevention, United States of America.

## Results

### Sociodemographic Characteristics of Study Participants

A total of 341 participants took part in the study, given a response rate of 100%. Participants’ modal age group was 25 to 44 years (49.27%). Most respondents were Muslim (85.92%) and of Hausa/Fulani ethnicity (79.77%), with 64.22% having at least secondary education. Majority (83.87%) lived within Bauchi metropolis. The remaining sociodemographic data are shown in Table 1.

**Table 1.** Sociodemographic Characteristics of the Respondents.

| Sociodemographic variable | Frequency | Percentage (%) |
| --- | --- | --- |
| **Age** |  |  |
| 18-24 | 56 | 16.42 |
| 25-44 | 168 | 49.27 |
| 45-64 | 98 | 28.74 |
| 65-85 | 19 | 05.57 |
| **Gender** |  |  |
| Male | 137 | 40.18 |
| Female | 204 | 59.82 |
| **Tribe** |  |  |
| Hausa/Fulani | 272 | 79.77 |
| Igbo | 8 | 02.34 |
| Yoruba | 6 | 01.76 |
| Others | 55 | 16.13 |
| **Religion** |  |  |
| Christianity | 48 | 14.08 |
| Islam | 293 | 85.92 |
| **Education** |  |  |
| No formal education | 75 | 22.00 |
| Primary education | 47 | 13.78 |
| Secondary education and above | 219 | 64.22 |
| **Occupation** |  |  |
| Public Servant | 102 | 29.92 |
| Self-employed | 118 | 34.60 |
| Unemployed | 121 | 35.48 |
| **Marital Status** |  |  |
| Currently Married | 258 | 75.66 |
| Not currently Married | 83 | 24.34 |
| **Place of residence** |  |  |
| Within Bauchi | 286 | 83.87 |
| Outside Bauchi | 55 | 16.13 |
| **Average family income (Naira)** |  |  |
| <50,000 | 122 | 35.78 |
| >50,000-<100,000 | 51 | 14.96 |
| ≥100,000 | 55 | 16.13 |
| Undeclared | 113 | 33.13 |
| \$1 equivalent to 324.2 Naira in 2018 | | |

Hypertension was the commonest morbidity among the participants (34.60%;118/341), followed by diabetes (7.62%; 26/341), with 5.28% (18/341) having both diabetes and hypertension. In the study, 291 participants had used herbal medicine before, resulting in a lifetime prevalence of 85.34%. Eighty participants were currently using herbal medicine (80/291; 27.49%), 88 (30.24%) had used it within the preceding twelve months, but only 14 (4.81%) had used it throughout the preceding twelve months. Ninety-eight (83.05%) of the 118 with hypertension and 22 (84.62%) of the 26 with diabetes used herbal medicine. Herbs were mostly used for treatment (248; 85.22%), and health promotion (102; 35.05%) among the participants. The commonest motivation for using herbs was their effectiveness (Table 2).

**Table 2.** Motivations for Using Herbal Medicine (n=291)

| Reason | Frequency | Percentage (%) |
| --- | --- | --- |
| It is effective | 123 | 42.27 |
| Better than nothing | 65 | 22.34 |
| It is easily available | 62 | 21.31 |
| It is natural medicine | 50 | 17.18 |
| Religious recommendation | 38 | 13.06 |
| It is cheap | 33 | 11.34 |

\*Multiple responses allowed

Participants classified the herbs they used as traditional medicine (90.72%; 264/291) and Islamic medicine (51.20%; 149/291).

Abdominal pain was the commonest conditions treated with herbal medicine (36.08%). About a sixth of the participants used herbal medicine for febrile illnesses (Table 3)

**Table 3.** Conditions Treated with Herbal Medicine.

| Condition | Frequency | Percentage (%) |
| --- | --- | --- |
| Abdominal pain | 105 | 36.08 |
| Hypertension | 43 | 14.78 |
| Febrile illness | 34 | 16.49 |
| Headache | 33 | 11.34 |
| Body pain | 32 | 11.00 |
| Sexual performance<br>enhancement | 21 | 7.22 |
| Pregnancy and labour | 17 | 5.84 |
| Cough | 14 | 4.81 |

\*Multiple responses allowed

Table 4 shows fifteen of the herbs commonly used by participants. Many of them are familiar, edible, local herbs.

**Table 4.** Herbs reported in the study.

| Herb |  |  | Frequency n<br>(%) |
| --- | --- | --- | --- |
| Local (Hausa) name | English name | Botanical name |  |
| Zogale | Moringa | *Moringa oleifera* | 174 (59.8) |
| Habatu-sawda | Black seed | *Nigella sativa* | 107 (36.8) |
| Zaytun | Olive oil | *Olea europaea* | 100 (34.4) |
| Tafarnuwa | Garlic | *Allium sativum* | 96 (33.0) |
| Ararabi | Frankincense | *Boswellia dalzielli* | 95 (32.6) |
| Hulba | Fenugreek | *Trigonella foenum-graecum* | 65 (22.3) |
| Lemun tsami | Lemon | *Citrus limon* | 56 (19.2) |
| Mankadai | Shea butter oil | *Vitellaria paradoxa* | 44 (15.1) |
| Citta | Ginger | *Zingiber officinale* | 42 (14.4) |
| Bagaruwa | Acacia | *Acacia nilotica* | 39 (13.4) |
| Namijin goro | Bitter kola | *Garcinia kola* | 31 (10.7) |
| Yayan magarya | Lote tree seed | *Ziziphus mauritiana* | 31 (10.7) |
| Dogoyaro | Neem | *Azadirachta indica* | 30 (10.3) |
| Ganyen zobo | Roselle | *Hibiscus sabdariffa* | 29 (10) |
| Kankana | Water melon | *Citrullus lanatus* | 29(10) |

### Association between Herbal Medicine Use and Sociodemographic Characteristics

Table 5 shows association between herbal medicine use and sociodemographic characteristics. Two sociodemographic factors – religion and place of residence – were significantly associated with herbal medicine use in the study population, with Muslims (P = 0.0005), and those living outside Bauchi (P= 0.01) more likely to use herbs.

**Table 5.**
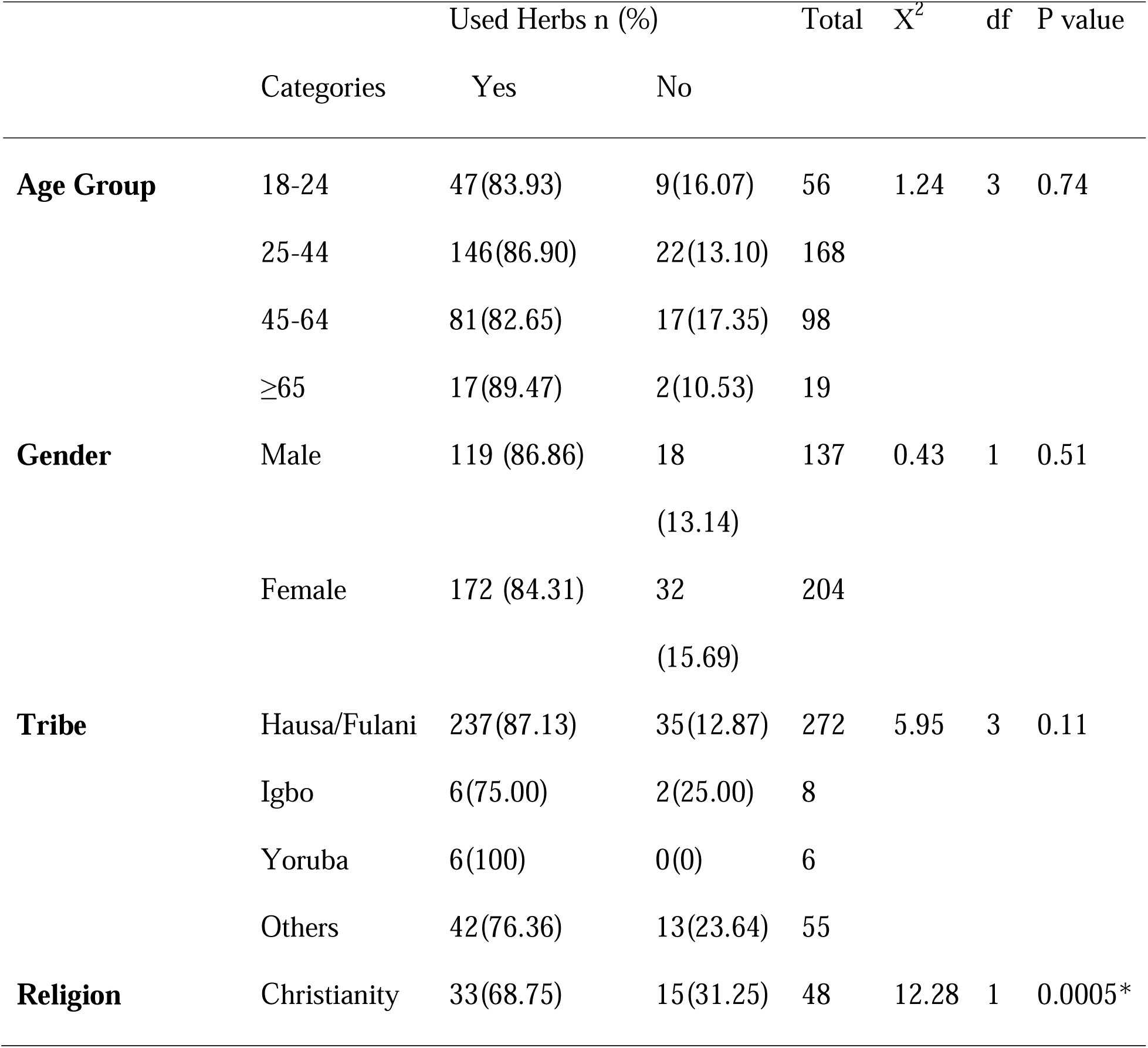
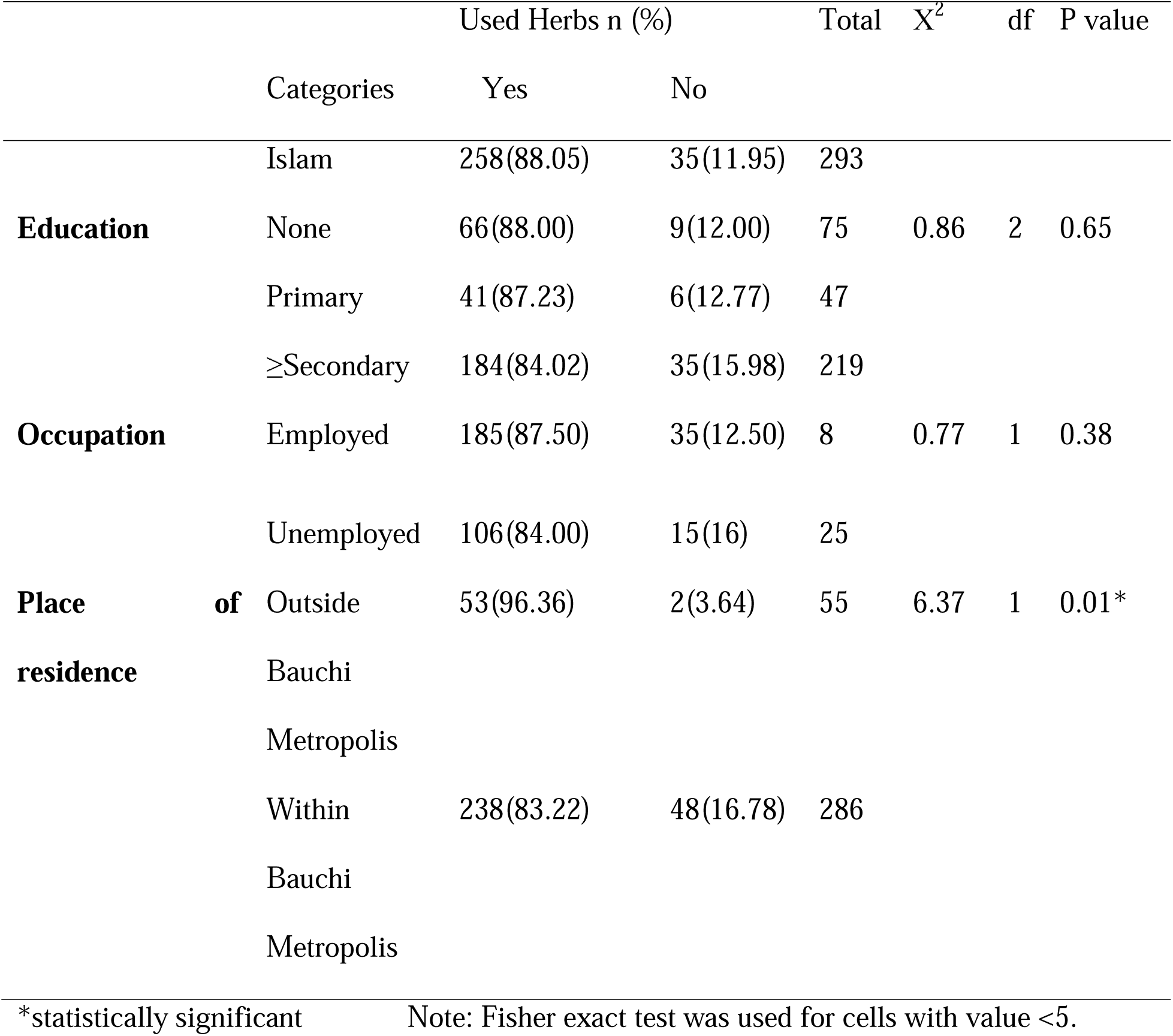
Association between herbal medicine use and sociodemographic characteristics.

Participants reported very few side effects (Table 6). The commonest was abdominal pain (4.12%)

**Table 6.**
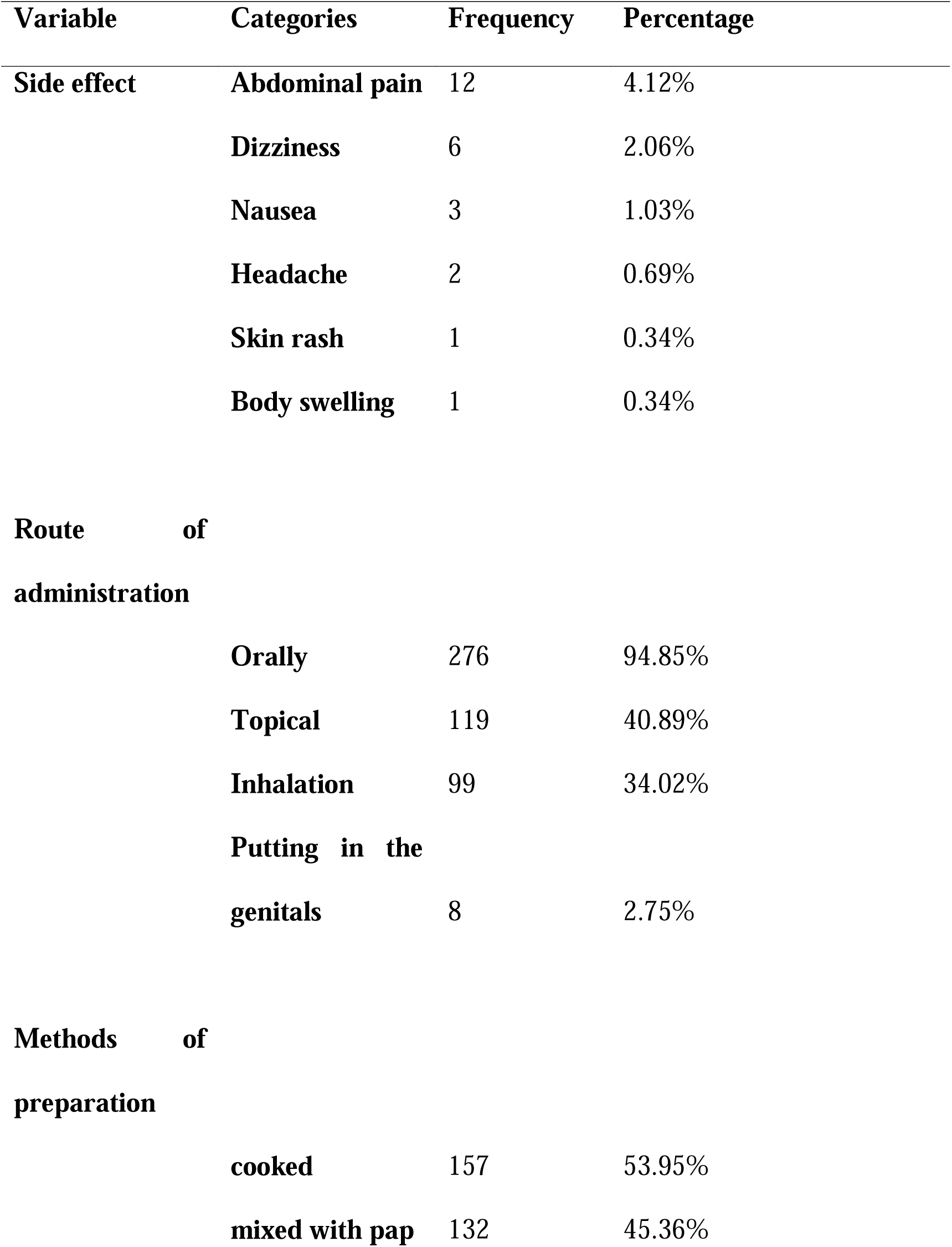
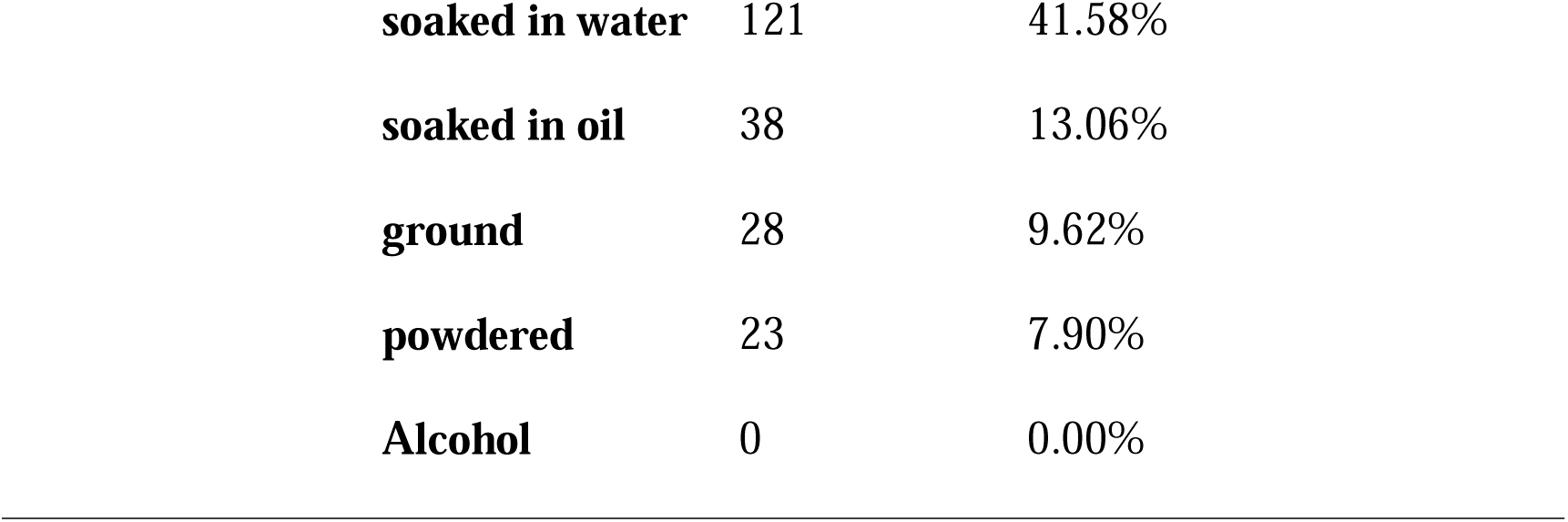
Side effect, route of administration and methods of preparation (n=291)

| **Variable** | **Categories** | **Frequency** | **Percentage** |
| --- | --- | --- | --- |
| **Side effect** | **Abdominal pain** | 12 | 4.12% |
|  | **Dizziness** | 6 | 2.06% |
|  | **Nausea** | 3 | 1.03% |
|  | **Headache** | 2 | 0.69% |
|  | **Skin rash** | 1 | 0.34% |
|  | **Body swelling** | 1 | 0.34% |
| **Route of administration** |  |  |  |
|  | **Orally** | 276 | 94.85% |
|  | **Topical** | 119 | 40.89% |
|  | **Inhalation** | 99 | 34.02% |
|  | **Putting in the genitals** | 8 | 2.75% |
| **Methods of preparation** |  |  |  |
|  | **cooked** | 157 | 53.95% |
|  | **mixed with pap** | 132 | 45.36% |

|  |  |  |
| --- | --- | --- |
| **soaked in water** | 121 | 41.58% |
| **soaked in oil** | 38 | 13.06% |
| **ground** | 28 | 9.62% |
| **powdered** | 23 | 7.90% |
| **Alcohol** | 0 | 0.00% |

The commonest route of administration was oral. Inhalation was reported by over a third of study participants (Table 6). Herbal medicines were prepared in different ways, mostly by cooking. However, participants did not report the use of alcohol as a solvent (Table 6).

Most participants who used herbal medicine obtained the products from herbal medical practitioners and herbal vendors (79.03%). Of note, doctors and other health workers very rarely were the source of herbal medicine (Table 7).

**Table 7:** Source of procurement of herbal medicine.

| **Source** | **Frequency** | **Percentage** |
| --- | --- | --- |
| **Herbal medical** |  |  |
| **practitioner** | 122 | 41.92% |
| **Herbal vendors** | 108 | 37.11% |
| **Self-made** | 79 | 27.15% |
| **Friends and family** | 61 | 20.96% |
| **Chemist/Pharmaceutical** |  |  |
| **shop** | 1 | 0.34% |
| **Other health workers** | 1 | 0.34% |
| **Doctors** | 0 | 0.00% |

Only 18.12 % of participants ever discussed herbal medicine use with their doctors (Table 8). The major reason cited for not discussing was that the doctors never asked (195/291; 81.59 %).

**Table 8:** Reasons for not discussing use of herbs with doctors.

| Reasons | Frequency | Percentage (%) |
| --- | --- | --- |
| Doctor did not ask | 195 | 81.59 |
| I was afraid of the doctor's response | 16 | 6.69 |
| I did not think it was important | 22 | 9.21 |
| There was no time to discuss | 2 | 0.84 |
| Others | 4 | 1.67 |

## DISCUSSION

The study aimed to determine characteristics of herbal medicine use among patients attending the general outpatient clinic of Abubakar Tafawa Balewa University Teaching Hospital. In all, 341 participants were enrolled. The modal age group was 25 –44 years (49.27%), with about 87% in that age range being users of herbal medicine (Table 1), suggesting the popularity of herbal medicine among the youth. Similarly, among those with at least secondary level of education, over 80% used herbal medicine. These findings indicate the popularity of herbal medicine among the young and the educated in Nigeria. Local studies corroborating these findings include Ahwinahwi et al. who reported that 81.78% of young undergraduates in Delta State used complementary and alternative medicine (CAM), with herbal medicine as the main CAM; and Osuchukwu et al in Calabar, where 75.5% of the respondents had at least secondary education.[4,8]

Most respondents were Muslims (85.92 %), the religion of the Hausa/Fulani tribe who constitute 79.77 % of the respondents. As such, religious influences are reflected in the prevalent pattern of herbal use. For instance, ‘Habatus-sawda’ (*Nigella sativa*) used commonly in this study is hardly reported in any study in the southern part of Nigeria. Its popularity among the study population is traceable to religious sources, where it is recommended as an elixir.[11,12] This religious influence is buttressed by the distinction participants made between traditional medicine (90.72%) and Islamic medicine (51.20%), otherwise known as Prophetic medicine. [13]

The lifetime prevalence of herbal medicine use in the study was 85.34%. This supports the estimate of the World Health Organization that about 80% of the population in Africa use traditional medicine.[1] Mbada reported a similar prevalence of 83.8% in Oyo State, though among rural farmers.[5] However, it appears that for the general population in our study, herbal medicine use was not a continuous practice, as only 4.81 % of the population continued to use herbal medicine throughout the year. This is in stark contrast to what was reported by Mbada et al. in South-West Nigeria, that as many as 96.8% of their respondents used herbs regularly at any time of the year. The occupation of the study participants in the latter study might explain this: most of the participants were farmers and used herbs mainly for relief of musculoskeletal pain, a constant result of their profession.

Our study found a very high prevalence of herbal medicine use among patients with hypertension (83%) and diabetes (85%), exceeding the prevalence reported in other studies in the southern part of the country. Among hypertensives in Ibadan, Osamor and Owumi found a prevalence of 29.1%, while Ogbera et al. reported a prevalence of 46% among diabetic patients in Lagos.[14,15] The few numbers of participants with hypertension and diabetes in our study may account for this discrepancy. Larger studies in Northern Nigeria among these two patient subgroups are recommended for better comparison.

Treatment was the main reason for use of herbal medicine by most respondents (85.22%), though a good proportion of the population also used herbs for health promotion (35.05%). Similar reasons were reported in other studies in Nigeria. In Calabar, Nigeria, Osuchukwu et al. found that the commonest reason why people used herbs was treatment, while in Uyo, South-South Nigeria, the commonest reason reported by Idung and his colleagues was health promotion and disease prevention.[4,6] Like in many previous studies, the commonest motivation for using herbs were effectiveness (42.27%), easy availability (21.31%), and naturalness (17.18%).[4,6,16,17]

The commonest conditions treated with herbal medicine in this study (Table 3) was abdominal pain. Meanwhile in Calabar, malaria and skin infection were the main conditions for herbal medicine use, and in Lagos, majority used herbs for ‘no specific reason’.[3,4] This variation in reasons for herbal medicine use reflects the different health priorities among different communities: health practitioners need to understand the health needs in their areas of practice.

The place of residence of respondent and their religion were significantly associated with use of herbal medicine (p=0.01 and p=0.0005 respectively) [Table 5]. Muslims were more likely to use herbs in this study. This was because of their double sources of herbal medicine: the Hausa/Fulani tradition and the influence of the aforementioned Islamic medicine, used by over 50% of those who used herbs. Religion was also significantly associated with herbal medicine use in Malaysia, a predominantly Muslim country.[18]

Place of residence was the second sociodemographic variable significantly associated with herbal medicine use, similar to the findings of Idung et al in Uyo.[6] This suggests that the environment could influence access to and use of herbs.

Despite the high prevalence of herbal medicine used and number of herbs used, very few side effects were reported. This is probably because many of the herbs, such as moringa and olive oil, are generally safe and edible.

The commonest route of administration of herbal medicine was oral, as in other studies.[19,20] However, the significant use of the nasal route (inhalation constituted 34.02 %) could exacerbate respiratory problems, especially for people with pre-existing respiratory diseases such as asthma and chronic obstructive pulmonary disease.[21]

Like in many African traditions, herbal medicine was prepared by cooking and soaking in water or oil (decoctions and infusions).[19,20] However, alcoholic solvents were not reported in this study most probably due to the aforementioned influence of the Islamic religion, which strictly forbids the use of intoxicants, even for medical purposes.[22]

The major sources of herbal medicine were outside the hospital: the herbal medical practitioners and the herbal vendors. Unlike in Norway, Germany, Japan and other countries, where doctors and other health workers receive training in herbal medicine and other CAMs, and offer them to interested patients or refer them to CAM practitioners, only one participant reported a health worker was the source of herbal medicine.[2,23,24] This reflects a dichotomy between orthodox medicine and traditional medicine in Nigeria, as against the integrative medicine model envisioned by the WHO.[1]

Less than a fifth of participants who used herbs (18.12 %) ever discussed their use of herbal medicine with doctors, and the chief reason for non-communication was that the doctors did not ask (81.59 %). Djuv et al in Norway similarly reported that only 15% of herb users discussed this with their general practitioners, and the major reason for this was because the doctor did not ask.[23]

Major limitations of the study are that it did not quantify the doses of herbal medicine used nor map specific herbs to particular ailments. Hopefully, future studies will focus on these.

The study reported that herbs commonly used by the respondents were often local, edible, familiar plants and that the principal motivation for use was assumed herbal efficiency. Unlike the studies in other parts of Nigeria, religion was a significant factor in the herbal medicine practice of Northern Nigeria. Finally, the study found that most patients source their herbal medicine from herbal medical practitioners and herbal vendors, and never discussed their use of herbal medicine with their doctors, suggesting CAM is yet to be integrated into mainstream medicine in Nigeria. With the high prevalence of herbal medicine use in Nigeria, it is recommended that the government increases research efforts into traditional medicine, in order to integrate it with orthodox medicine as recommended by the WHO.

## Data Availability

All data produced are available online at:
DOI: 10.6084/m9.figshare.25062332

